# Statistical Design and Analysis of Diagnostic Tests for Mutating Viruses

**DOI:** 10.1101/2021.04.07.21254917

**Authors:** Yang Han, Yujia Sun, Jason C. Hsu, Thomas House, Nick Gent, Ian Hall

## Abstract

As the SARS-CoV-2 virus mutates, mutations harboured in patients become increasingly diverse. Patients classified into two strains may have overlapping non-variant-defining mutations.

Mutation calling by sequencing is relative to a *reference genome*. As SARS-CoV-2 mutates, tracking emerging mutant strains may become increasingly problematic if the reference genome remains Wuhan-Hu-1, because the comparison then becomes *indirect*: current dominant strain relative to Wuhan-Hu-1 versus emerging strain relative to Wuhan-Hu-1.

The original Thermo Fisher’s TaqPath PCR test, on which the UK has standardized national testing of SARS-CoV-2 primarily, targets Wuhan-Hu-1. PCR targets appear readily updated, as TaqPath 2.0 now targets both currently known and future SARS-CoV-2 mutations, probing the N gene and ORF1ab but not the S gene, with 8 probes instead of the original 3 probes. Going forward, our statistical method can more *directly* compare current wildtype versus emerging mutants, since our new method can use any pair of probes updated to probe the current wildtype and anticipated mutations.

The fact that patients harbour mixtures of mutations allows our statistical methods to potentially catch emerging mutants. Given a PCR test which targets the current dominant strain (current wildtype), our statistical method can potentially *directly* differentiate the current wildtype from an emerging strain.

## 1 Introduction

SARS-CoV-2 is a single stranded RNA virus that mutates. These mutations give rise to so-called variants of concern (VoC). A variant of concern (VoC) is one that increases transmissibility, causes the disease to be more severe or diagnostic detection to fail, or reduces effectiveness of treatments or vaccines. In the UK, the current variants of concern are B.1.1.7 (from South East England), B.1.351 (from South Africa), P.1 (from Brazil) and B.1.617.2 with its sub-lineages AY.1, AY.2, and AY.3 (from India) [6]. In the U.S., B.1.1.7 (Alpha), {B.1.351, B.1.351.2, B.1.351.3} (Beta), {B.1.617.2, AY.1, AY.2, AY.3} (Delta), and {P.1, P.1.1, P.1.2} (Gamma) variants are classified as variants of concern. This article proposes a PCR-based statistical method that can track emerging variants to guide sequencing efforts.

PCRs are popular diagnostic tests for SARS-CoV-2. In the UK, the TaqPath PCR test is the principal analytic platform for Pillar 2 which consists of key workers in the NHS, social care and other sectors. Pillar 2 is also the diagnostic pathway for community cases. Such tests typically have multiple probes targeting different regions of the SARS-CoV-2 genome to bind to. Outcome of a PCR test is a Cycle threshold (Ct) for each target region, the thermal cycle number at which the fluorescent signal exceeds that of the background. Ct is a semi-quantitative value that can broadly categorize whether the amount of the target genome sequence in the sample is high or low. While a low Ct indicates substantial presence of the target, a high Ct may be due to either the amount of the target in the sample being low, or a mutation has caused a decrease in binding of the probe to the target (as the probe is built on the original genome sequence of SARS-CoV-2).

Sequencing is the gold standard for tracking virus mutations. But PCR testing is magnitudes cheaper than sequencing. Analysis of real data sets containing PCR as well as sequencing results reveal that even PCR testing can discover emerging mutant of the SARS-CoV-2 virus. The statistical method we have developed can separate an emerging mutant strain from wildtype using one pair of PCR probes, any pair. It does a reasonable job if wildtype and the emerging mutant strain are sufficiently “apart” in lineages, but has limitation otherwise, as we will describe.

Explanation of why our PCR-based method can work comes from the observation that patients tend to harbour *mixtures* of mutation. That is, each patient may contain mutations observable in more than one strain of the SARS-CoV-2 virus (classified according to Pangolin lineage [9]). We share this observation with the scientific community, as it may have implication beyond our immediate goal.

### 2 Idea of An Emerging Variant Monitoring System

Our premise is, at any given time, there is a dominant (majority) strain (call it wildtype), and *potentially* an emerging (minority) mutant strain that might, in time, become dominant. (While there are almost surely other mutant strains, we assume their prevalence are quite low.^1^) Our goal is to develop a PCR-based monitoring system, to detect signal of an emerging mutant strain before it becomes dominant, to focus sequencing effort on.

In this monitoring situation, one does not know whether there is a mutant strain emerging. If there is a subpopulation forming a cluster of new mutations, the hope is that PCR testing can detect the cluster before sequencing can. As one does not know the emerging mutation sequences in advance, there is no data available to train PCR Ct counts on, which cases should be classified as wildtype and which as mutants. Thus, in the classical statistical learning sense, our setting is *unsupervised* learning (with two unknown clusters).

However, our simple statistical method changes the problem formulation in an unconventional way: we *target* one PCR probe as the classifying probe, and we choose another PCR probe to *baseline* the target. Then we use a statistical technique to choose a classification cut-point for the targeting probe. Taking some liberty with terminologies, we *train* our algorithm based on PCR data only, and then *validate* it by estimating its *sensitivity*, the probability that a true mutant is called as such, using either available sequencing results, or by statistical modelling.

#### 2.1 Variant Definitions

Variants are designated according to Pangolin phylogenetic classification (e.g., B.1.1.7), Public Health England (PHE)’s VOC/VOI year-date nomenclature (e.g., VOC-20DEC-01), and WHO’s Greek alphabet scheme (e.g., Alpha).

WHO’s variant Alpha, Delta, … naming scheme has become popular. The Pangolin phylogenetic tree classification scheme is more informative, in giving a sense of how close or far apart the different variants are. B.1 strains are (presumably) closer to each other than to the P.1 strain, for example.

Our data comes from PHE, so sequence-calling of case as of a particular variant is made according to PHE’s variant-defining mutations. While each patient may harbour dozens of mutations, s/he is said to be infected by a particular strain if her/his mutations contain a variant-defining subset.

For example, [4] defines an infected person as a **confirmed** B.1.1.7 (VOC-20DEC-01, Alpha) if the persons has all 13 *variant-defining* mutations in Table 1, a **probable** B.1.1.7 case if the person does not have all of those 13 mutations but at least 5 of them. The 69-70del (deletion of amino acids 69 and 70) made the world aware of the mutation of S gene back in September 2020. Most B.1.1.7 patients have the 69-70del which causes S gene target failure (SGTF) but the 69-70del is in fact not included in the B.1.1.7 variant definition. Table 1 also lists the 29 additional mutations have been observed from 21 January 2021 to 20 April 2021 in persons classified as B.1.1.7-infected, as reported in Table 11 of [8].

**Table 1:** B.1.1.7 Variant-defining and additional mutations, see [4] and [8]

| Variant-defining mutations | Additional Mutations |  |
| --- | --- | --- |
| N501Y | L18F | L455F |
| A570D | Q677H | G476S |
| P681H | P384L | Q677R |
| T716I | A701V | K147E |
| S982A | F490S | A684S |
| D1118H | S255F | P9S |
| T1001I | P9L | K150R |
| A1708D | K458N | Q493R |
| I2230T | S494P | A684V |
| Q27 | P384S | H146Y |
| R52I | A684T | Q493K |
| Y73C | E484K | S12P |
| Confirmed if case has all 13 mutations | D253G |  |

As another example, [4] defines an infected person as a Confirmed B.1.617.2 (VOC-21APR-02) if that person has at least 8 out of 12 variant-defining mutations listed in Table 2, which also lists the 29 additional mutations in the spike gene of the B.1.617.2 (Delta) variant observed as of 15 June 2021, as reported in Table 13 of [5].

**Table 2:** B.1.617.2 Variant-defining and additional mutations, see [4] and [5]

| Variant-defining mutations | Additional mutations |  |
| --- | --- | --- |
| T19R | G142D | L244S |
| D63G | P681L | T716I |
| T478K | P251L | L18F |
| R203M | G446V | S477I |
| P681R | V483F | R158G |
| L452R | S494L | P479S |
| S26L | K417N | K444N |
| D950N | S494A | Q677H |
| I82T | P384S | D405Y |
| V82A | R683Q | D215G |
| T120I | R683L | S255F |
| D377Y | E484A | E484Q |
| Confirmed if case | V503I | G142A |
| has at least 8 | R346G | Q321L |
| out of 12 mutations | A701V |  |

It seems to us plausible that some infected patients harbour **mixtures** of mutations observed in more than one strain. What we observe in our data supports this theory^2^. Our statistical method works by detecting and separating mixtures in mutations in infected persons.

#### 2.2 A Uniform PCR platform for Contact Tracing in UK

To track how the CoV-2 virus is spreading, and mutating, uniformity of test results is important.

For uniformity of PCR test results, the UK decided early on to standardize Covid-19 infection testing on Thermo Fisher’s TaqPath PCR test, processed by Lighthouse laboratories. A Lighthouse laboratory is a high throughput facility that is dedicated to COVID-19 testing for UK’s National Testing Programme.

Sampling and testing of potential infected persons utilize Tier 2 dedicated professional staff of the Contact Tracing Advisory System (CTAS), focusing on Pillar 2 workers. Any person who has been in contact with a confirmed infected person is labelled a “Contact” in the database. A person becomes a “Case” if that person later tests positive by PCR.

Pillar 2 consist of critical key workers in the NHS, social care and other sectors. Rationale for focusing on them is they are front line health care workers frequently coming into contact with infected persons. Besides efficient tracking of spread of Covid-19, PCR testing surprisingly offers a potential early warning system for emergence of mutated CoV-2 variant. Sequencing can then be focused on where the emergent variant clusters, and containment measure can be instituted if needed.

#### 2.3 Data Sets

Our June 2021 data set is Variants of Concern under investigation data created from 1 November 2020 up to 21 June 2021 [3]. Total sample size *n* = 1,048,575. Among those sequenced, *n* = 22,479 are called as B.1.1.7, and *n* = 6,406 are called B.1.617.2. Among the B.1.1.7 cases, *n* = 15,680 have complete non-zero Ct values for the N gene and ORF1ab, but only 58 have Ct for the S gene. Among the B.1.617.2, *n* = 5,144 have complete non-zero Ct values for the N gene and ORF1ab, of which *n* = 5,124 have Ct for the S gene as well. There were other variants called, but few, see Table 3. So this June 2021 data set is a good example of our premise, that at a snapshot in time, there is a dominant strain (B.1.1.7 in June 2021) and an emerging more transmissible strain (B.1.617.2).

**Table 3:** Variants with highest prevalence in June 2021 data set

| PHE VOC/VOI | Pangolin | WHO | No. of cases | First Identified |
| --- | --- | --- | --- | --- |
| 20DEC-01 | B.1.1.7 | Alpha | 22479 | UK (Kent) |
| 20DEC-02 | B.1.351 | Beta | 84 | South Africa |
| 21JAN-02 | P.1 | Gamma | 23 | Brazil* |
| 21APR-02 | B.1.617.2* | Delta | 6440 | India |

\*and Japan, \*with AY1 and AY2

We also have two earlier data sets. Our November 2020 data set Variants of Concern under investigation data created from 28 May 2020 to 2 November 2020 [2]. Total sample size *n* = 1, 048, 575. Among those samples, *n* = 500,440 are Covid-19 Positive by PCR or culturing. Among those confirmed Covid-19 Positive samples, *n* = 309, 139 have complete Ct values for the N gene, the S gene, and ORF1ab. As the B.1.1.7 strain was first identified on 20 September 2020, this data set likely contained both patients infected by the original SARS-CoV-2 strain as well as those infected by the B.1.1.7 variant.

Our February 2021 data set is Variants of Concern under investigation data created from 1 November 2020 up to 3 February 2021, from the COG-UK data set, PHE Second Generation Surveillance System and the PHE Rapid Investigation Team Kent investigation [3]. (Sequencing results are available on only a small percentage of the positive cases as PCR testing capacity far exceeds the sequencing capability.) Total sample size *n* = 7,582,696. Among the *n* = 2,646,907 confirmed Covid-19 cases (called *Positive* by PCR or culturing), *n* = 2,561,279 are missing sequencing test results. Among those sequenced, *n* = 38,612 are called as B.1.1.7, *n* = 116 are called as B.1.351, with *n* = 46,900 called as Wildtype. Confirmation of being B.1.1.7 is by phylogenetic tree matching analysis with the B.1.1.7 strain. Wildtype is defined as sequenced samples other than B.1.1.7 and B.1.351. Within those called B.1.1.7 by sequencing, *n* = 34,018 are further classified as Confirmed B.1.1.7, *n* = 4,536 as Probable B.1.1.7, and *n* = 58 as Low-quality Genome. Among the Confirmed and Probable B.1.1.7, only *n* = 76 have complete Ct values for the N gene, the S gene, and ORF1ab.

## 3 Methods

We initially analysed our first, the November 2020 data set, by simply plotting the distribution of the S gene’s Ct, to see if the density indicates it is a mixture of distributions, one for patients infected by (what was then considered) wildtype, one or more others infected by mutant strains, see Figure 1. Neither the histogram nor its density version indicates anything definitive. So apparently PCR data tends to be noisy, there is too much person-to-person variability in the S gene’s Ct within those infected by wildtype, and within those infected by some mutant strain, that their distributions overlap too much for us to tell which patients are infected by wildtype and which are infected by some mutated variant(s).

**Figure 1:**
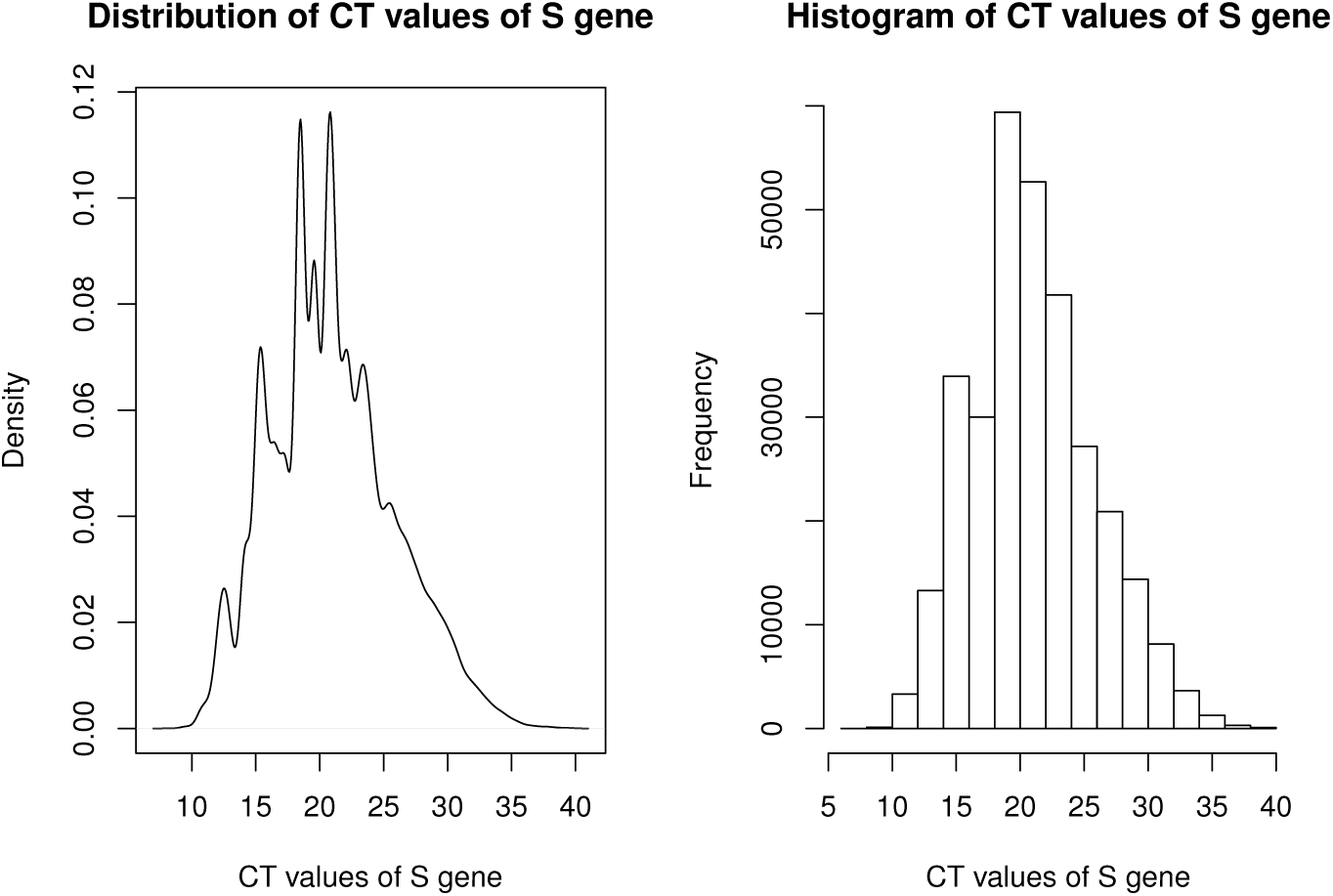
Distributions of CT values of S gene in the November 2020 data set.

We thus examine the possibility of controlling for person-to-person variability using one or more covariates of the S gene’s, specifically Ct’s of the N gene and ORF1ab. We view them simply as variables that might be used to “baseline” each patient, a *covariate* in statistical terminology.

### 3.1 The Conditional Residual Statistical (CRS) Method

Conditioning is a tried and true principle of statistical inference.

Our statistical method, called the Conditional Residual Statistical (CRS) method, finds a controlling variable which suitably reduces a target gene’s Ct variability conditionally given the controlling variable’s value, then calculates the predicted value for the target gene’s Ct *disregarding potential mutation*. With our premise that patients contain mixtures of mutations, assuming wildtype still dominates, the method calls a patient as infected by wildtype if its predicted target value is in the dominant cluster, and calls a patient as infected by a mutant strain if its predicted target value is outside the dominant cluster. The conditioning technique we use is linear regression.

#### 3.1.1 Before the B.1.617.2 strain emerged

Using the November 2020 data set as an example, for each patient, Ct’s of the N gene and ORF1ab may be correlated with the Ct of the S gene (high or low in the same direction), so we might use the Ct of the N gene or Ct of ORF1ab or both to baseline each patient. Conditioning on Ct of ORF1ab turns out to work well.

Denoting *log*(*Ct*) of the S gene as *log*(*S*) and *log*(*Ct*) of ORF1ab as *log*(*ORF*1*ab*), as previously described, [7] trained the CRS method by regressing *log*(*S*) on *log*(*ORF*1*ab*) based on the November 2020 data set. Raw value of the S gene’s Ct is then replaced by its residual, its observed S gene Ct value minus its predicted S gene Ct value (given the corresponding Ct value of the conditioning variable). Call them S conditional residuals. Figure 2 shows the S conditional residual plot, which indicates it is a mixture of two densities, a dominant one concentrated on the left, and a more diffused one on the right.

**Figure 2:**
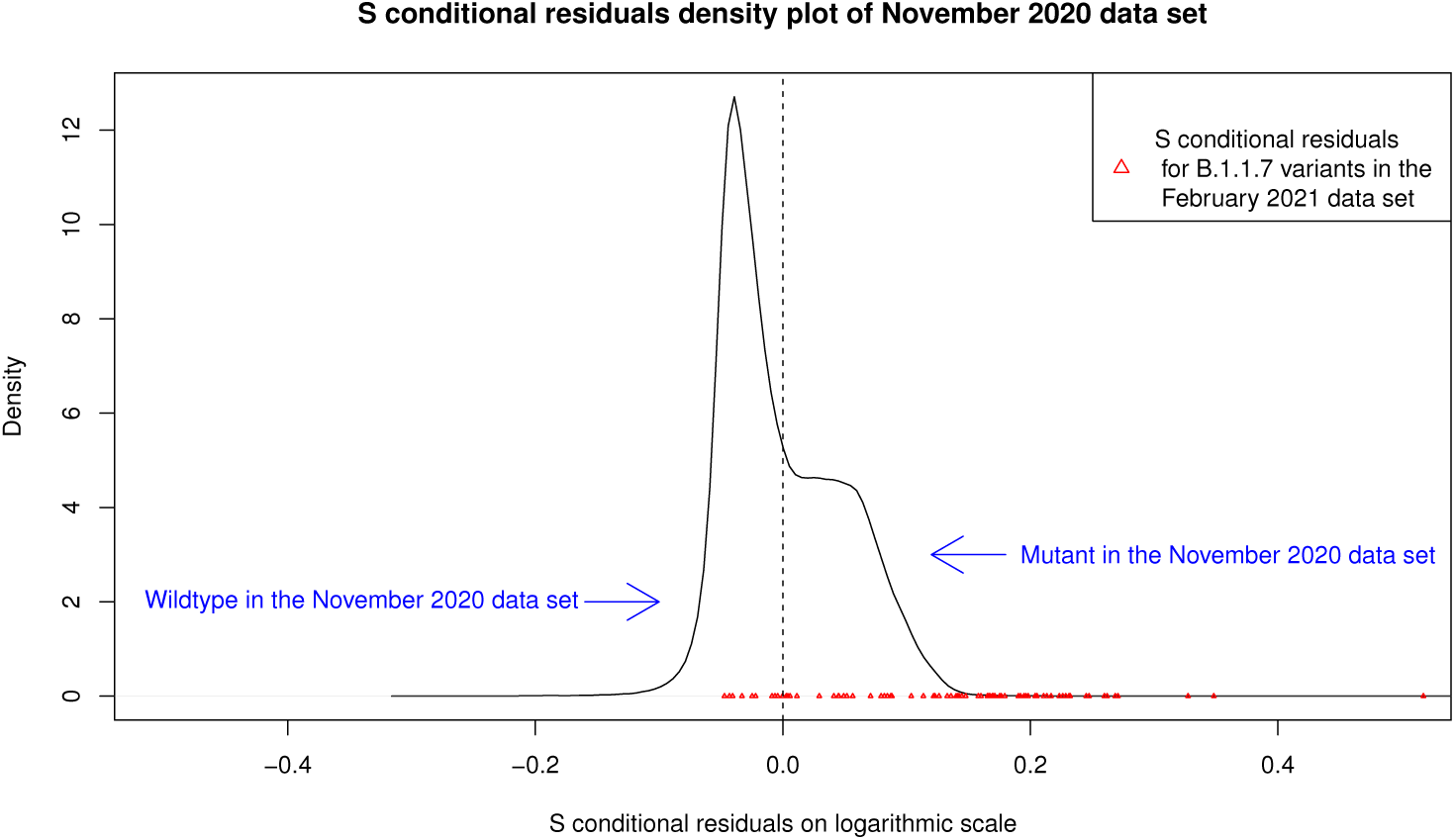
Training on the November 2020 data set validating with the February 2021 data set

One challenge of applying the CRS method is to decide on a cut-point to divide the clusters, to call a case as wildtype if its residual is on one side of the cut-point and call the case mutated if its residual is on the other side. For this November 2020 data, [7] simply chose zero to be the cut-point, since zero is one measure of the “center” of the residuals (average of the residuals is always zero), and zero seems a reasonable cut-point looking at Figure 2.

[7] then tested the CRS method using confirmed and probable B.1.1.7 cases (confirmed by sequencing) in the (separate) February 2021 data set, calling cases to the left of zero as wildtype, and cases to the right of zero as mutated. Represented by the red triangles in Figure 2, out of the 76 confirmed and probable B.1.1.7 cases, 66 (about 87%) have positive S-residuals while 10 (about 13%) have negative S-residuals. So, empirically, *sensitivity* of the CRS method, the probability that a B.1.1.7 is called as such is about 87%.

After [7] was written, we have added a statistical modelling technique to the CRS method, to help guide cut-point selection, as well as to estimate sensitivity. Figure 3 shows the result of *modelling* the mixture density as a mixture of two Normal densities statistically, using the `mixtools` R package. Looking at this figure, one might set a cut-point slightly to the left of zero but we keep it at zero for simplicity.

**Figure 3:**
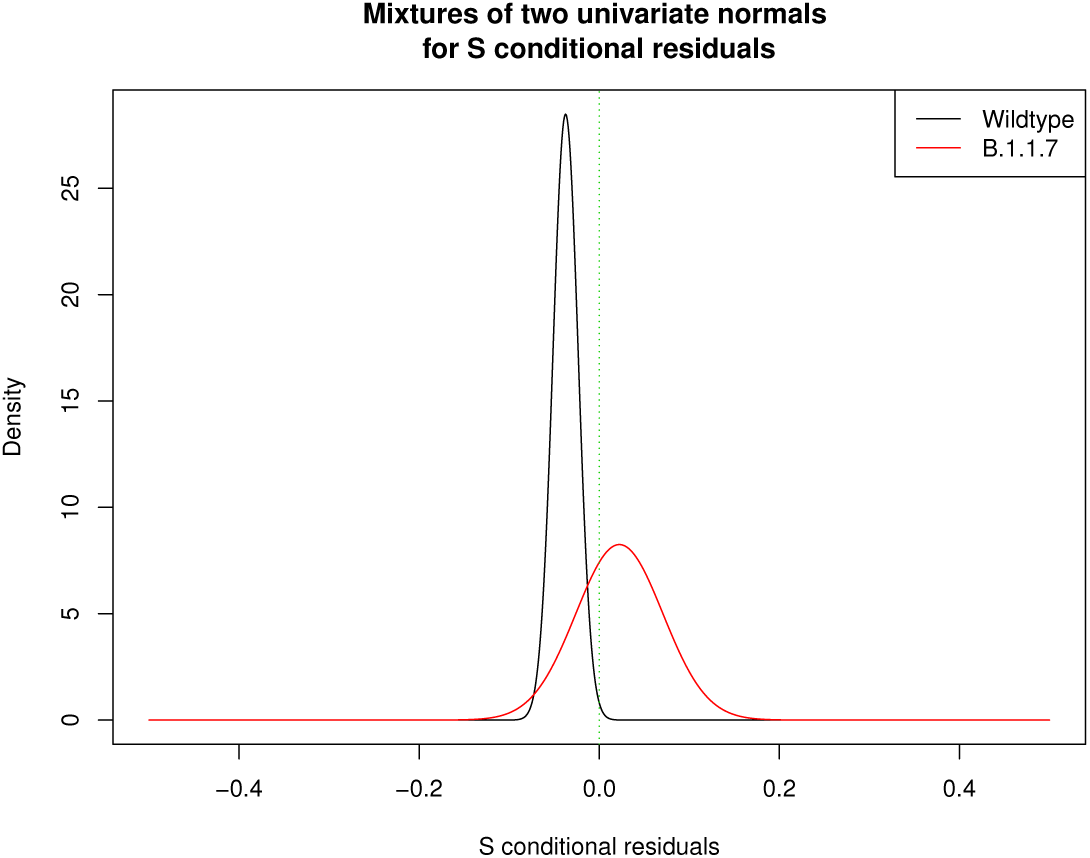
Modelling the November 2020 mixture densities

Estimated mean and standard deviation of wildtype are −0.037 and 0.014 respectively, while estimated mean and standard deviation of B.1.1.7 are 0.022 and 0.048 respectively. Based on the modelled B.1.1.7 density, *sensitivity* of CRS, the probability that a B.1.1.7 called by sequencing is called as such by CRS, is calculated to be about 68%, which is conservative compared to the empirically observed 87%.

With the modelled wildtype density almost entirely to the left of zero in Figure 3, modelling would predict that almost no wildtype would be called B.1.1.7. However, empirically only 17,581 out of the 27,269 sequenced wildtypes, about 65%, are correctly called as wildtype by CRS. So modelling seems overly optimistic in terms of *specificity*, the probability that a true wildtype is called as such by CRS. One possible explanation is that “wildtype” has changed from November 2020 to February 2021.

For monitoring emerging variants, to be able to estimate sensitivity is more important than to estimate specificity.

#### 3.1.2 After emergence of the B.1.617.2 strain

Applying our CRS method to the June 2021 data set confirms its potential but also reveals its limitation. To illustrate that our method can use any pair of probes, we consider using probes for the N gene and ORF1ab, not using probe for the S gene.

Figure 4 shows the residual density plot of log(ORF1ab) regressed on log(N), from which it is difficult to discern whether it is a mixture of densities or not. This contrast to Figure 3 puzzled us, since we know among the sequenced cases in the June 2021 data set, the 15,680 B.1.1.7 cases and the 5,144 B.1.617.2 cases dominate, and we expect to see two bumps.

**Figure 4:**
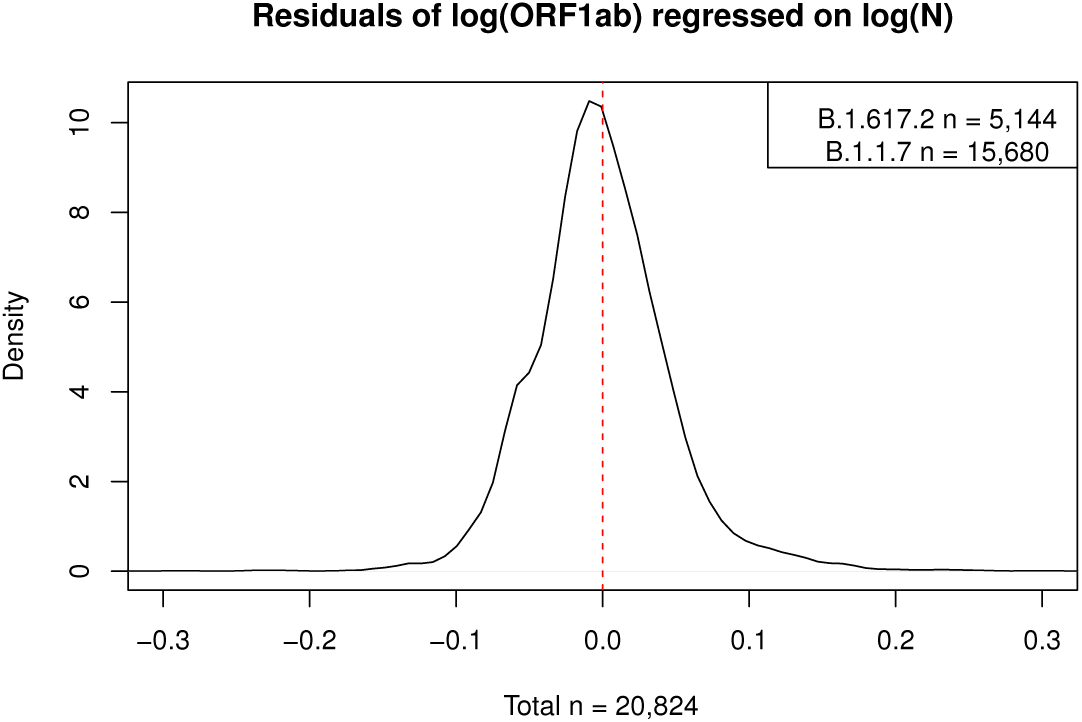
Residual density plot from the June 2021 data set does not clearly show two bumps.

We tried applying `mixtools` to model the June 2021 data set as a mixture of two Normal densities, but `mixtools` (which is based on the EM algorithm) essentially failed, giving rather variable results depending on the initial means we specify.

With plenty of B.1.1.7 and B.1.617.2 cases in the June 2021 data set, we made empirical density plots of them as shown in Figure 5, which clearly shows two densities. Reasons for the difficulty of modelling them as two Normal densities may be closeness of the two peaks, and skewness of the density of B.1.1.7, with a longer tail to the right. Zero would not work well as a cut-point, with the two peaks at or to the left of zero.^3^

**Figure 5:**
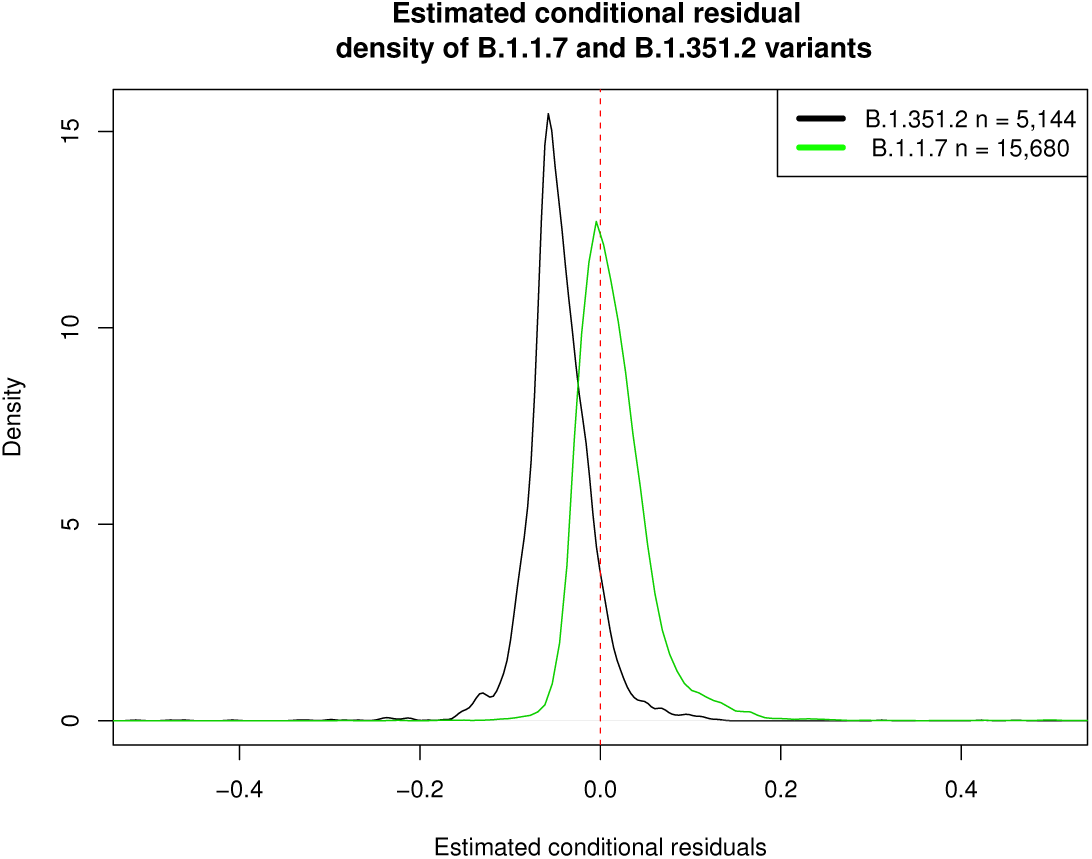
The two estimated densities are too close to be separated.

We further investigated, if wildtype and emerging mutant are relatively close in mutations, how prevalent does the mutant strain need to be before empirical (non-modelling) conditional residual plot can detect it. With wildype being B.1.1.7, Figure 6 shows CRS may need B.1.617.2 to becomes as prevalent as B.1.1.7, in order to pick up B.1.617.2.

**Figure 6:**
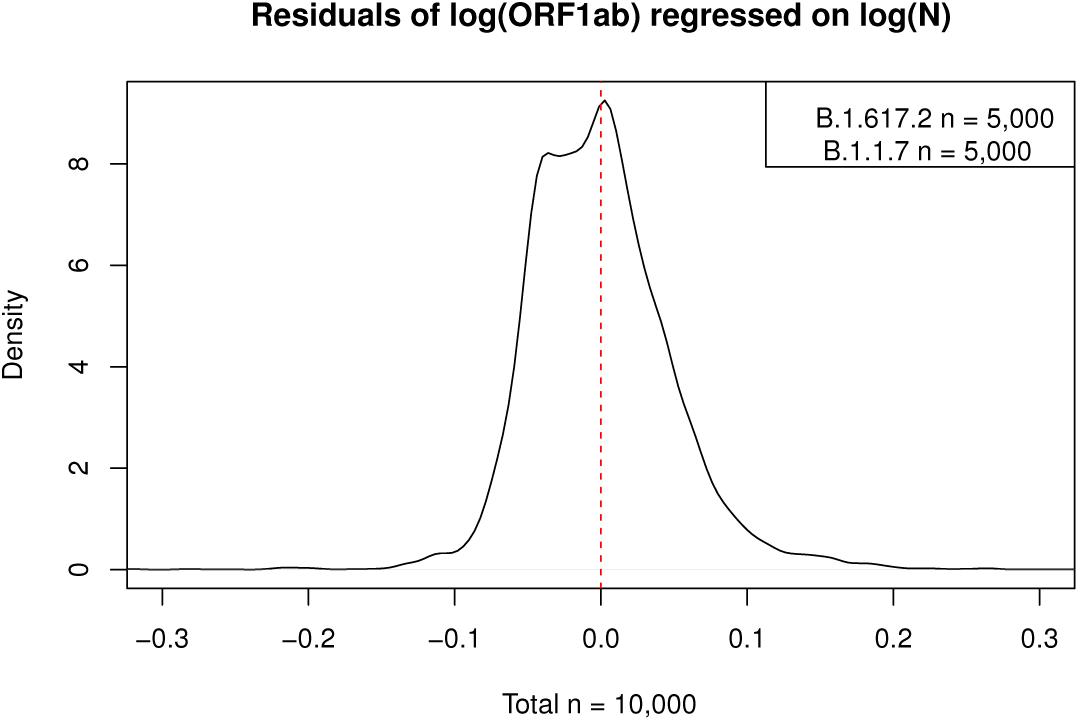
Mutant prevalence needs to be high to be detected if lineages are close.

To summarize, requirements for the CRS method to be an effective emerging variant monitoring system include

1. Mutations between wildtype and the emerging mutant strain are sufficiently apart to cause differential bindings by PCR probes;
2. Mixture density of the conditional residuals can be successfully de-mixed by some tool.

## 4 Monitoring Mutations by Observing Mixtures

WHO’s variant Alpha, Detla, … naming scheme has become popular in the media. WHO’s naming scheme gives the impression that variants are disjoint. In fact each strain contains a mixture of mutations.

The Pangolin scheme [9] is more realistic, classifying an infected person as an B.1.617.2 (Delta) rather than a B.1.1.7 (Alpha) if that person’s mutations’ phylogenetic distance is closer to the variant-defining mutations for B.1.617.2 than the variant-defining mutations for B.1.1.7 or other variants. Figure 3a of [8] shows the phylogenetic tree placement of the B.1.617.1 (VUI-21APR-0, Kappa), B.1.617.2, and B.1.617.3 variants.

Mutation calling by sequencing is relative to a *reference genome*, which by default is Wuhan-Hu-1. As the SARS-CoV-2 virus continues to mutate, tracking emerging mutant strains will become problematic if the reference genome stays Wuhan-Hu-1, because the comparisons are *indirect* : B.1.1.7 relative to Wuhan-Hu-1 compared to B.1.617.2 relative to Wuhan-Hu-1, for example. Our CRS method can directly differentiate the current wildtype from an emerging strain using a PCR test targeting both currently known and future SARS-CoV-2 mutations.

## 5 Comparison with Machine Learning

For comparison, we applied the unsupervised machine learning technique of K-Means clustering to the ORF1ab and the N gene Ct counts, asking it to find two clusters. The K-Means clustering completely fails to separate variants.

Figure 7 shows the result of 2-Means Clustering, which basically puts cases with high N and ORF1ab Ct counts in the B.1.617.2 cluster, and puts cases with low N and ORF1ab Ct counts in the B.1.1.7 cluster.

**Figure 7:**
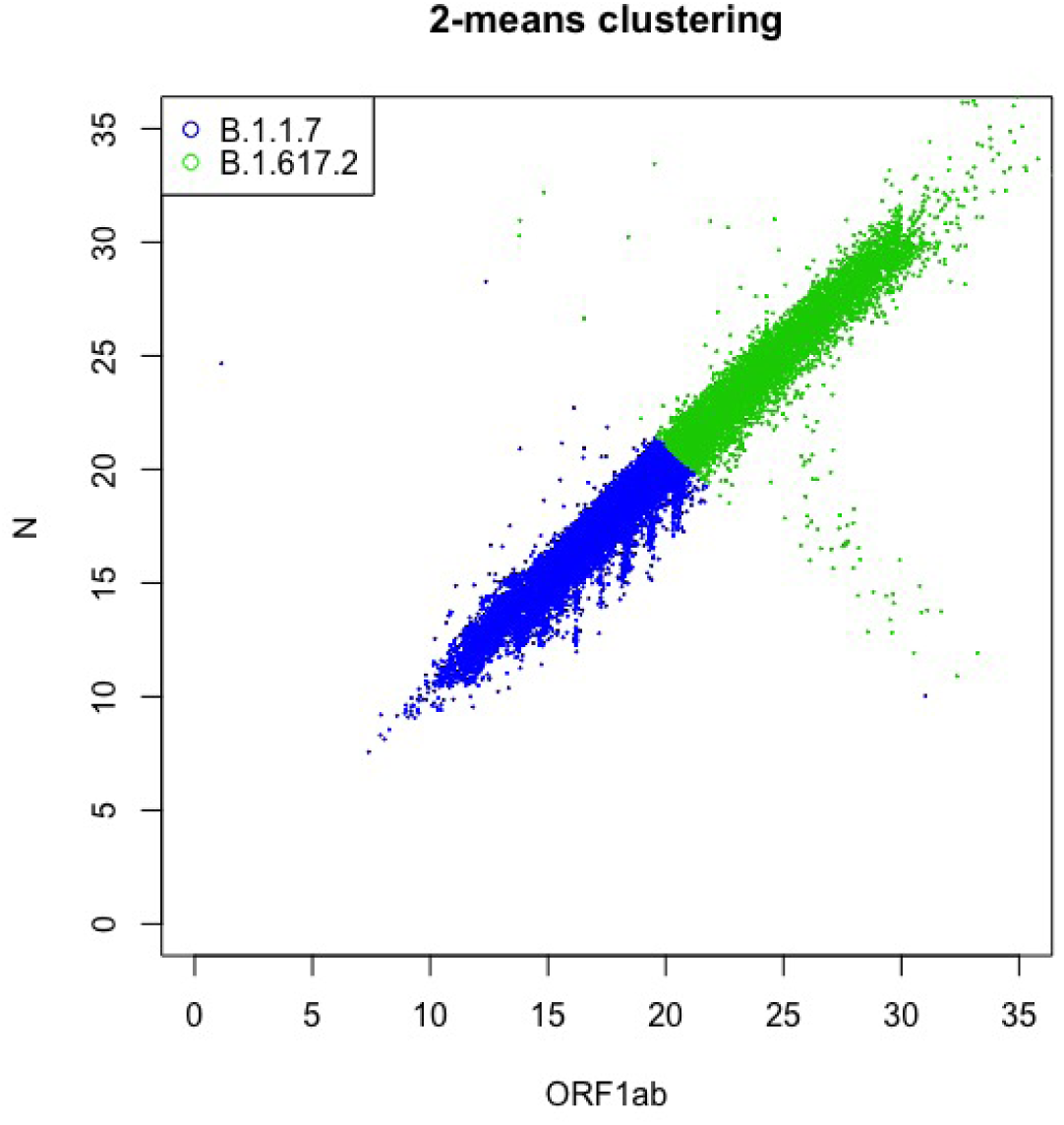
The two clusters found for the June 2021 data by 2-Means Clustering

Figure 8 reveals the true identity of the cases, which shows 2-Means Clustering completely fails to differentiate between B.1.617.2 from B.1.1.7. The reason for the failure of 2-Means clustering is that N and ORF1ab Ct counts are highly correlated, and whether they are high or low largely reflects noise in the Ct counts. The reason our statistical method can catch emerging mutants (with some limitation) is our method makes innovative use of domain knowledge. We use the biological knowledge that different samples have different amount of viral materials, and the statistical knowledge that conditioning can remove such sample-to-sample variation.

**Figure 8:**
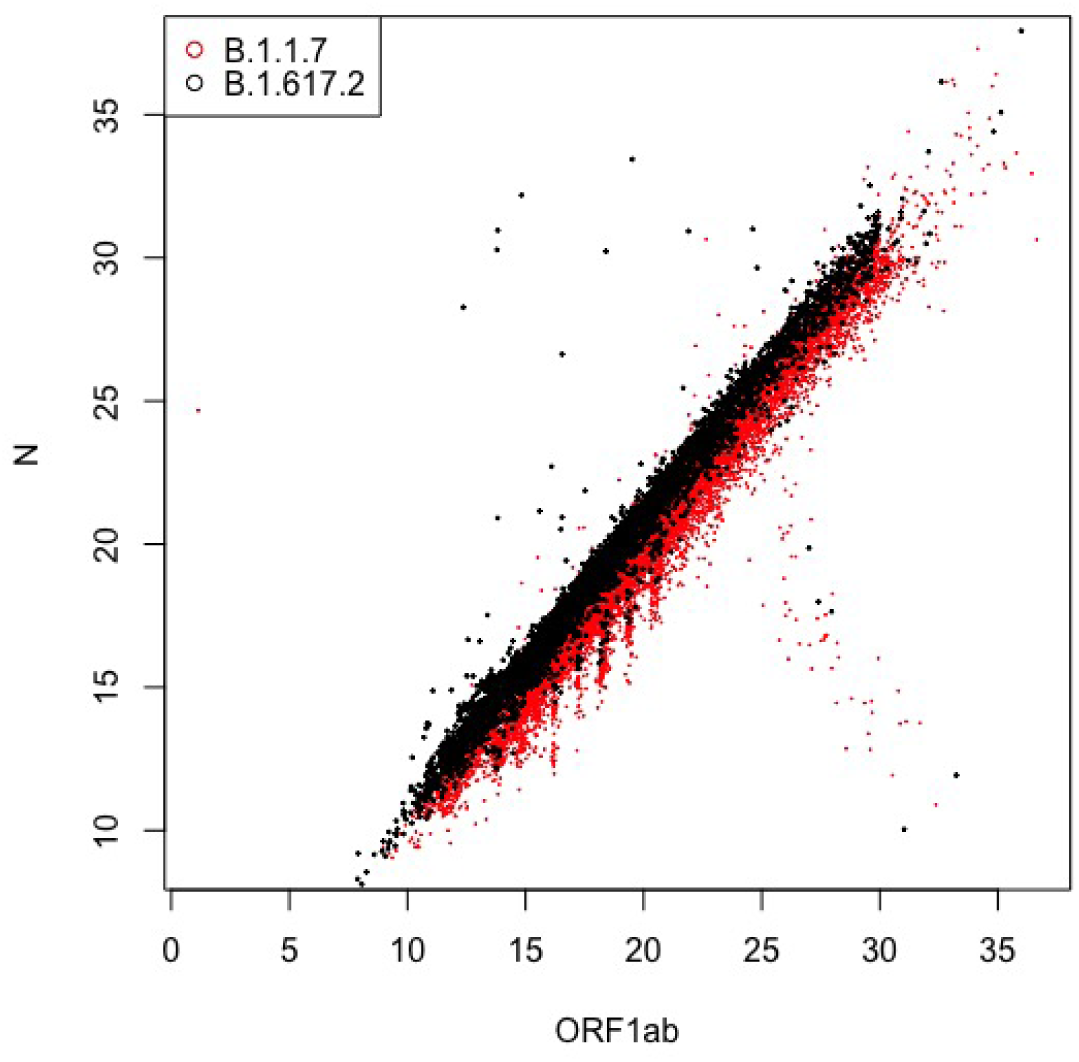
True identity of B.1.1.7 and B.1.617.2 cases in the June 2021 data

## 6 Recommended Design Principles

We suggest the following principles for designing PCR for SARS-CoV-2:

1. Target multiple genes and/or open reading frames, both known mutation sites, as well as potential (future) mutation sites.
2. Choose probes that are biologically as functionally independent as possible Our rationales are as follows.

### Principle 1

Designing probes for PCR traditionally is based on the sequence of a fixed (not mutating) strain. The challenge of designing probes for SARS-CoV-2 is that if the probes are based on the dominant strain at the moment, that dominant strain may not be the most prevalent for very long. So perhaps one should design probes that can capture the current dominant strain(s) and future mutations. In order to provide increased confidence in COVID-19 test results as SARS-CoV-2 continues to mutate, For example, Thermo Fisher introduced TaqPath 2.0 in July 2021, which has 8 targets instead of 3 in the original TaqPath. Whereas the original TaqPath has one probe in the S gene, one in the N gene, and one in ORF1ab, TaqPath 2.0 has 3 probes in ORF1a, 2 probes in ORF1b, and 3 probes in the N gene.

### Principle 2

Technically, making the probes statistically independent to classify patients tends to work better when the probes are more biologically functionally independent. This is because, with more bio-logical independence, there is a higher chance that a suitable internal Control variable can be found to act as the baseline for the target gene.

## 7 Final Remark

Both PCR probe designers as well as sequencing-based mutation callers need to continuously update their *reference genome*, to reflect what is the most prevalent strain at the current time. For both, it would be useful if the scientific community can get together periodically to agree on a current SARS-CoV-2 reference genome. This would promote uniformity of reporting of PCR positivity rates. Subsequent mutation-calling by sequencing then will be relative to the current consensus reference genome, as opposed to relative to the increasingly distant Wuhan-Hu-1.

## Data Availability

No publicly accessible data is available for sharing.

## Footnotes

1 This assumption is bore out by what we observed in the June 2021 data set, described in Section 2.3.

2 Our ^2^On 13 July 2021, researchers in Belgium report on the case of a 90-year-old woman who was both B.1.1.7 and B.1.351 [1].

3 Note that the B.1.1.7 density in Figure 5 is that of residuals of ORF1ab, whereas the B.1.1.7 density in Figure 3 was of S gene residuals.

## References

[1] European Society Of Clinical Microbiology And Infectious Diseases. 90-year-old woman infected with UK and South African COVID-19 variants at the same time. News Release, July 2021. https://www.eurekalert.org/news-releases/526106.

[2] Public Health England, London, UK. Public Health England, 2020.

[3] Public Health England, London, UK. Public Health England, 2021.

[4] Public Health England (PHE). PHE genomics variant definitions. https://github. com/phe-genomics/variant_definitions, 2021. [Online; accessed 12-September-2021].

[5] Public Health England (PHE). SARS-CoV-2 variants of concern and variants under investigation in England-technical briefing 16. https://assets.publishing.service.gov.uk/government/uploads/system/uploads/attachment_data/file/1001359/ Variants_of_Concern_VOC_Technical_Briefing_16.pdf, 2021. [Online; accessed 12-September-2021].

[6] Public Health England (PHE). SARS-CoV-2 variants of concern and variants under investigation in England-technical briefing 22. https://assets.publishing.service.gov.uk/government/uploads/system/uploads/attachment_data/file/1014926/Technical_Briefing_22_21_09_02.pdf, 2021. [Online; accessed 12-September-2021].

[7] Yang Han, Yujia Sun, Jason C Hsu, Thomas House, Nick Gent, and Ian Hall. Statistical design and analysis of PCR tests for fast mutating viruses. medRxiv, 2021. https://www.medrxiv.org/content/early/2021/04/09/2021.04.07.21254917.

[8] Public Health England (PHE). SARS-CoV-2 variants of concern and variants under investigation in England-technical briefing 9. https://assets.publishing.service.gov.uk/government/uploads/system/uploads/attachment_data/file/979818/Variants_of_Concern_VOC_Technical_Briefing_9_England.pdf, 2021. [Online; accessed 12-September-2021].

[9] A Rambaut, E.C. Holmes, and Á. O’Toole et al. A dynamic nomenclature proposal for SARS-CoV-2 lineages to assist genomic epidemiology. Nature Microbiology, 5:1403, 2020.

